# Impact of Community Masking on SARS-CoV-2 Transmission in Ontario after Adjustment for Differential Testing by Age and Sex

**DOI:** 10.1101/2023.07.26.23293155

**Authors:** Amy Peng, Savana Bosco, Alison Simmons, Ashleigh R. Tuite, David N. Fisman

**Author notes:** The research was supported by a grant to DNF from the Canadians Institutes for Health Research (2019 COVID-19 rapid researching funding OV4-170360). ART was employed by the Public Health Agency of Canada when the research was conducted. The work does not represent the views of the Public Health Agency of Canada. **Address reprint requests and correspondence to:** David Fisman, MD MPH FRCP(C) Room 686, 155 College Street, Toronto, Ontario, M5T 3M7.

## Abstract

**Background:** Use of masks and respirators for prevention of respiratory infectious disease transmission is not new, but has proven controversial, and even politically polarizing during the SARS-CoV-2 pandemic. In the Canadian province of Ontario, mask mandates were introduced by the 34 regional health authorities in an irregular fashion from June to September 2020, creating a quasi-experiment that can be used to evaluate impact of community mask mandates. Ontario SARS-CoV-2 case counts were strongly biased by testing focussed on long-term care facilities and healthcare workers. We developed a simple regression-based test-adjustment method that allowed us to adjust cases for undertesting by age and gender. We used this test-adjusted time series to evaluate mask mandate effectiveness.

**Methods:** We evaluated the effect of masking using count-based regression models that allowed adjustment for age, sex, public health region and time trends with either reported (unadjusted) cases, or testing-adjusted case counts, as dependent variables. Mask mandates were assumed to take effect in the week after their introduction. Model based estimates of effectiveness were used to estimate the fraction of SARS-CoV-2 cases, severe outcomes, and costs, averted by mask mandates.

**Results:** Models that used unadjusted cases as dependent variable identified protective effects of masking (effectiveness 15-42%), though effectiveness was variably statistically significant, depending on model choice. Mask effectiveness in models predicting test-adjusted case counts was substantially higher, ranging from 49% (44- 53%) to 73% (48-86%) depending on model choice. Effectiveness was greater in women than men (P = 0.016), and in urban health units as compared to rural units (P < 0.001). The prevented fraction associated with mask mandates was 46% (41-51%), averting approximately 290,000 clinical cases, averting 3008 deaths and loss of 29,038 QALY. Costs averted represented $CDN 610 million in economic wealth.

**Conclusions:** Lack of adjustment for SARS-CoV-2 undertesting in younger individuals and males generated biased estimates of infection risk and obscures the impact of public health preventive measures. After adjustment for under-testing, the effectiveness of mask mandates emerges as substantial, and robust regardless of model choice. Mask mandates saved substantial numbers of lives, and prevented economic costs, during the SARS-CoV-2 pandemic in Ontario, Canada.

## Introduction

The use of masks and respirators for prevention of respiratory disease transmission is not new (1–3). However, during the SARS-CoV-2 pandemic the use of these tools has proven controversial (4), and even politically polarizing (5). Supporters and opponents of community masking can point to a mixed evidentiary database when it comes to the impact of masking for reducing disease risk (6). Challenges in assessment of mask effectiveness in community settings include potential confounding in observational studies, unintended crossover and contamination in randomized trials (7), and the difficulty in teasing apart the bidirectional impacts of masking (prevention of inhalation of viral particles as well as reduction in infectious aerosol generation in infective individuals) (8).

Masking directives were typically issued at times of increased community risk, which may again serve to obscure the impact of masking. Lastly, studies that rely on surveillance data, and evaluate masking using quasi-experimental designs, may be limited by the differential and targeted use of PCR testing in different groups in the population. For example, we previously found that testing in the Canadian province of Ontario tended to be heavily targeted towards women aged 80 and over, who constitute the majority population in the province’s nursing homes (9). We found that adjustment for differential testing by age and sex resulted in a different picture of infection risk by age, with infection risk strongly concentrated in younger age groups once differential testing was accounted for.

In the Canadian province of Ontario, there was no provincial masking directive introduced in the summer of 2020. Rather, individual public health regions introduced mask mandates for indoor public settings between June and September 2020, with variable timing. This created an ideal quasi-experiment that permitted evaluation of the effects of community-level masking mandates on disease incidence. An econometric analysis was performed on these data by Karaivanov et al., who found that community masking likely had an efficacy of 22% in reducing SARS-CoV-2 transmission in Ontario (10). However, we hypothesized that the true impact of masking might be obscured by differential testing of the Ontario population by age and sex, with the impact of masking in younger age groups obscured by under-testing.

Our primary objectives were to evaluate the impact of community mask mandates in the period from March to December 2020 in the province of Ontario, using both reported SARS-CoV-2 case time series, and using time series adjusted for differential testing. A secondary objective was to estimate the SARS-CoV-2 prevented fraction for community masking in Ontario during this time period.

## Methods

### Data Sources

We evaluated masking impact using population-based SARS-CoV-2 infection data from the Ontario Case Management System, a data system used by Ontario public health units for public health management of notifiable diseases. The period of interest was from March 20, 2020, when community transmission of SARS-CoV-2 had clearly begun in Ontario, to December 8, 2020. Our reason for ending our analysis in early December 2020 was 3-fold: vaccination against SARS-CoV-2 was initiated in mid-December 2020 (11, 12); more centralized public health guidance for the province around SARS-CoV-2 came into force in December 2020 (13); and December 2020 marked the appearance of the first SARS-CoV-2 variants of concern with the N501Y mutation circulating in Ontario (14). Case count data were available as weekly time series, with counts stratified by 10-year age category, reported case gender (dichotomized as female/non-female), and public health unit (9). Data on weekly PCR test counts for SARS-CoV-2 by 10-year age category, reported case gender and public health unit were derived from the Ontario Laboratory Information System (9), and population denominators were obtained from Statistics Canada (15).

Testing rates were evaluated graphically and statistically using negative binomial regression. As highest test rates were seen in females aged 80 and over, we used meta-regression-based methods to adjust case counts in other age and gender groups for under-testing, estimating the case rates that would have been expected if these groups were tested at the same rate as females aged 80 and over. This method is described in detail elsewhere (9), but briefly requires that a standardized infection ratio (SIR), and standardized testing ratio (STR), be estimated weekly, by public health unit, for each age- and gender-group, with incidence and testing rates in females aged 80 and over used in the denominator of these ratios. It is then possible to create age- and gender-specific meta-regression models using log-transformed of *i* age and gender groups, E(ln(SIRi)) = α + ý(ln(STRi)). As ln(STRi) is zero when a given age and gender group is tested at the same rate as females aged 80 and over, the model intercept α can be interpreted as the standardized infection ratio that would be expected in the presence of equal testing. This SIR can then be multiplied by observed infection incidence in females aged 80 and over to generate an estimate of test-adjusted incidence. Weekly test-adjusted case estimates for each public health unit were generated in this way, and used as exposures in models as described below.

### Mask Effectiveness

We evaluated mask effectiveness through construction of negative binomial regression models with public health unit populations used as offsets. Using two different dependent variables: reported weekly case counts, and test-adjusted weekly case counts. We fit initial models that incorporated linear, quadratic and cubic time trends, age group (modeled as an ordinal variable), gender and provincial reopening stages for each health unit. As SARS-CoV-2 cases declined over the summer of 2020, the government employed a regional, staged approach to reopening, with progressively increasing venue occupancy and gathering sizes, depending on disease activity at the local public health unit level (16). The effects of staged reopening were considered to have taken effect one week after reopening. We then added community masking mandates to these base models, with masking mandates were assumed to take effect one week after introduction. We evaluated the effect of mask mandates based both on the P-value for the model coefficient and change in model fit as assessed via likelihood ratio test.

In our primary analysis (Model 1), we fit negative binomial regression models with public health units treated as indicator (dummy) variables. We added community masking to models and evaluated change in model fit based on the likelihood ratio test, as well as the P-value for the masking coefficient. Incidence rate ratios were estimated for masking, with confidence intervals adjusted for clustering by public health unit.

We evaluated the robustness of this finding in sensitivity analyses, where we repeated our analysis using alternate modeling approaches, including using cross-sectional time series negative binomial (panel data) approach, with public health units treated as fixed effects and confidence intervals estimated via bootstrapping (Model 2), as well as a hierarchical generalized linear modeling approach, with time series nested within public health units, which were treated as random effects (Model 3). For Model 3, a Poisson family and log link were used with the gllamm command in Stata 15 (17) as gllamm does not allow the use of the negative binomial family.

### Prevented Fraction

We estimated the prevented fraction of test-adjusted infections between first introduction of mask mandates on June 12, 2020 and December 8, 2020 based on Model 1 predictions with masking, and by generating Model 1 predictions with the community masking variable set to zero; we also estimated prevented fractions and 95% confidence intervals using the punaf command in Stata 15 (18). We estimated the health gains associated with cases averted through masking by applying Ontario- derived age-specific case-fatality, hospitalization and ICU admission risk estimates as in (19). For deaths averted, we assigned gains in quality-adjusted life-years (QALY) based on the approach of Briggs and Kirwin (20, 21). The economic costs averted due to hospitalizations and ICU admissions averted were assigned based on estimates from the Canadian Institute for Health Information (22); we assigned an economic value of $30,000 to each QALY gained based on the approach of Kirwin et al.(21). Maps were created using QGIS (23), while other statistical analyses were conducted in Stata version 15 (24). Aggregate data files needed for replication of results are available at https://figshare.com/articles/dataset/Data_for_Fisman_et_al_Test-Adjusted_Incidence_of_COVID-19_in_Ontario/14036528. The study was approved by the Research Ethics Board of the University of Toronto (protocol number 00044787).

## Results

There was a significant upward log-linear trend in testing rates over time (relative increase 4.30% per week, 95% CI 4.20-4.40%) (**Figure 1A**). Test rates in females were higher than in males (IRR 1.624, 95% CI 1.590 to 1.660), and test rates in those aged 80 and over were higher than in younger individuals (IRR 1.670, 95% CI 1.613 to 1.730) during the time period under study. In models in which each age and gender group was treated as a unique category, women aged 80 and over were tested at significantly higher rates than all other groups except females aged 20-49, in whom test rates were not significantly different (**Figure 1B**). As such, we used females aged 80 and over as our referent group for calculation of standardized testing and infection ratios, and for calculation of test-adjusted case counts.

**Figure 1.**
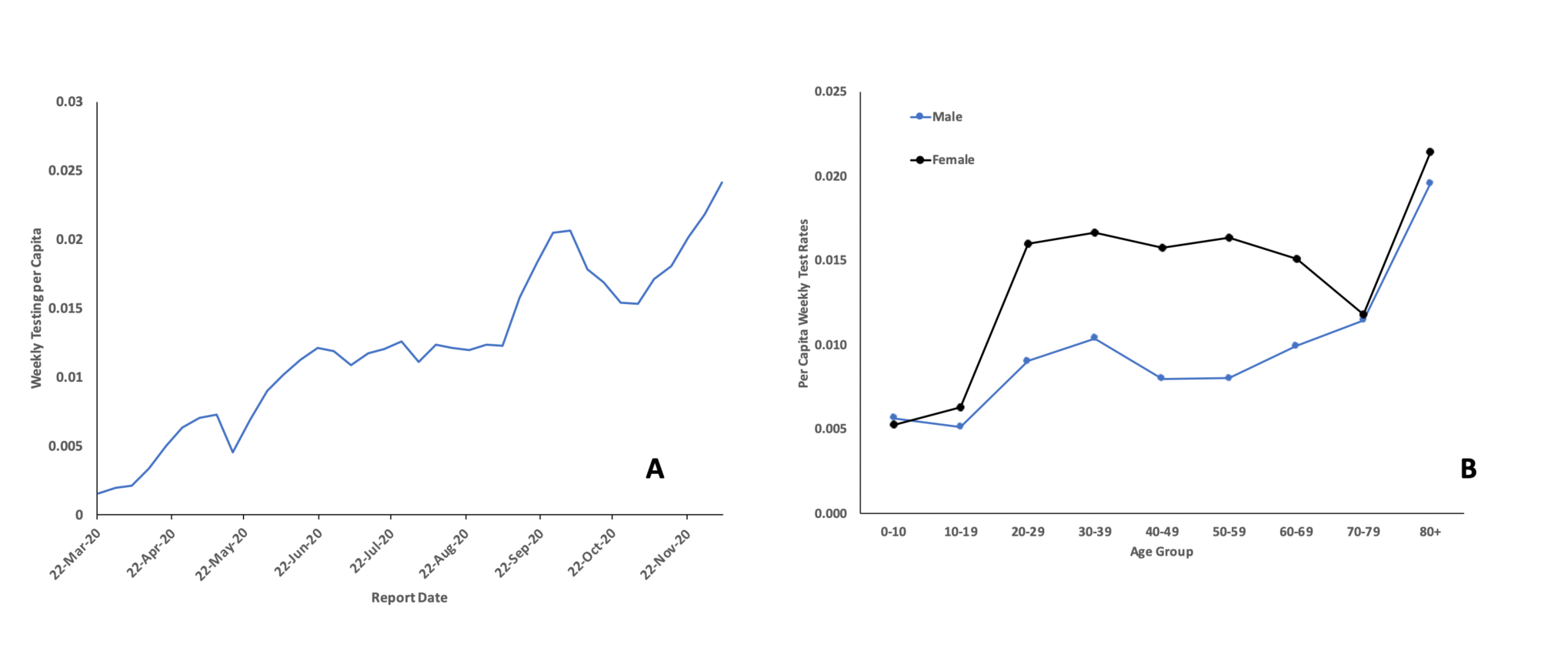
Trends in Testing for SARS-CoV-2 in Ontario, Canada, 2020. Weekly per capita tests are presented on the Y-axis in panels A and B. In panel A, test report date is presented on the X-axis. Testing increased at an average rate of 4.3% (95% CI 4.2% to 4.4%) from March to December 2020. Panel B presents weekly per capita tests by age group (X-axis) and sex. Females testing rates are plotted in black; blue curve represents testing in males.

The epidemic curve for SARS-CoV-2 in Ontario, and the epidemic curve created by adjusting for under-testing by age and sex, is presented in **Figure 2**. While the reported epidemic curve demonstrated highest incidence in autumn 2020, test-adjustment revealed a larger spring 2020 wave, with a smaller autumn wave.

**Figure 2.**
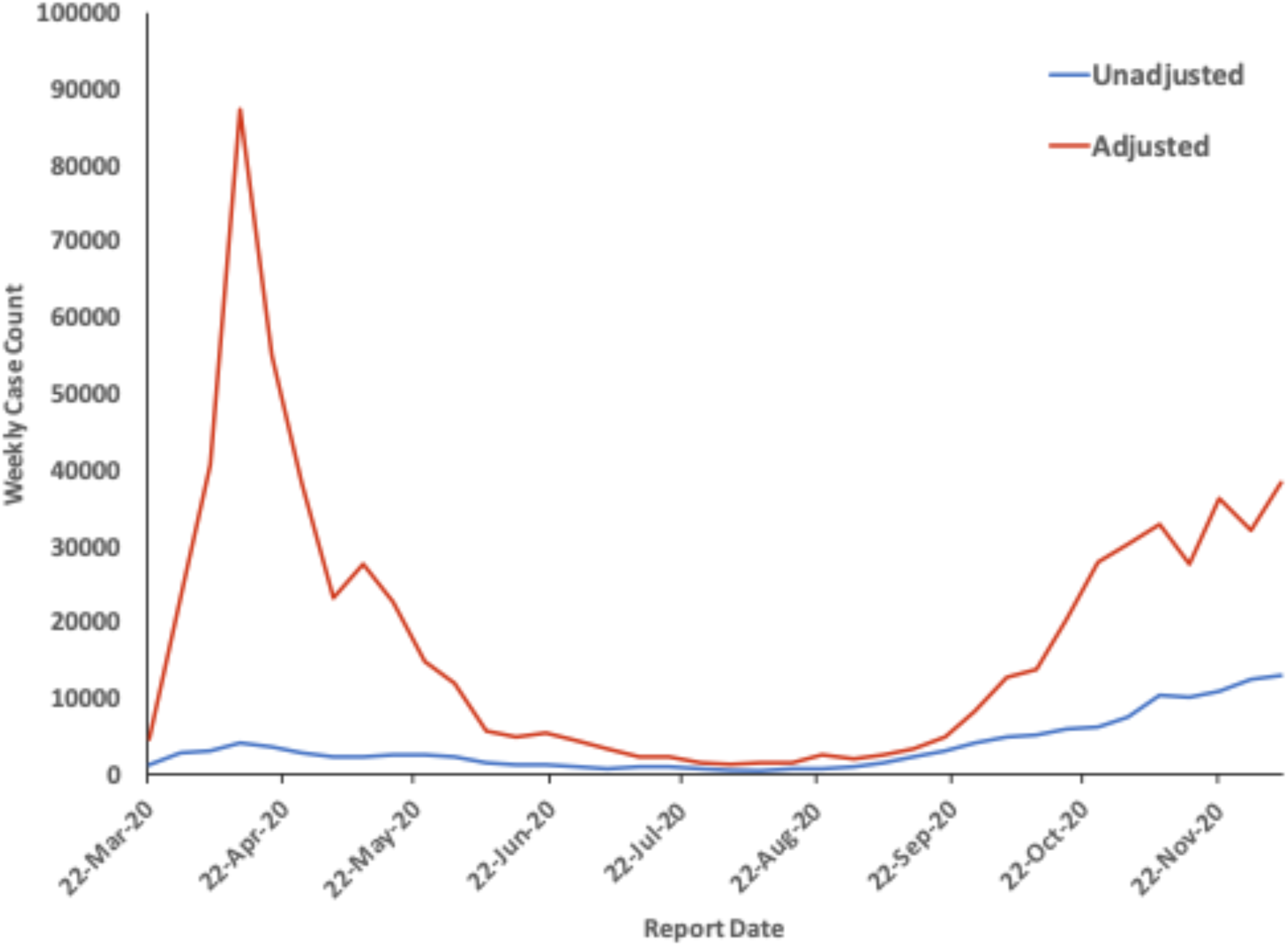
Ontario SARS-CoV-2 Epidemic Curve in 2020 With and Without Test Adjustment. Weekly case counts are presented on the Y-axis; test report dates are presented on the X-axis. The blue curve shows reported case counts without adjustment for under- testing. The red curve shows expected case counts if all age and sex groups were tested with the same intensity as women aged 80 and over.

Introduction of indoor mask mandates by individual public health units began on June 12, 2020 (Wellington-Dufferin-Guelph), with the Chatham-Kent public health unit the last to introduce a mask mandate (September 14, 2020) (**Figure 3**).

**Figure 3.**
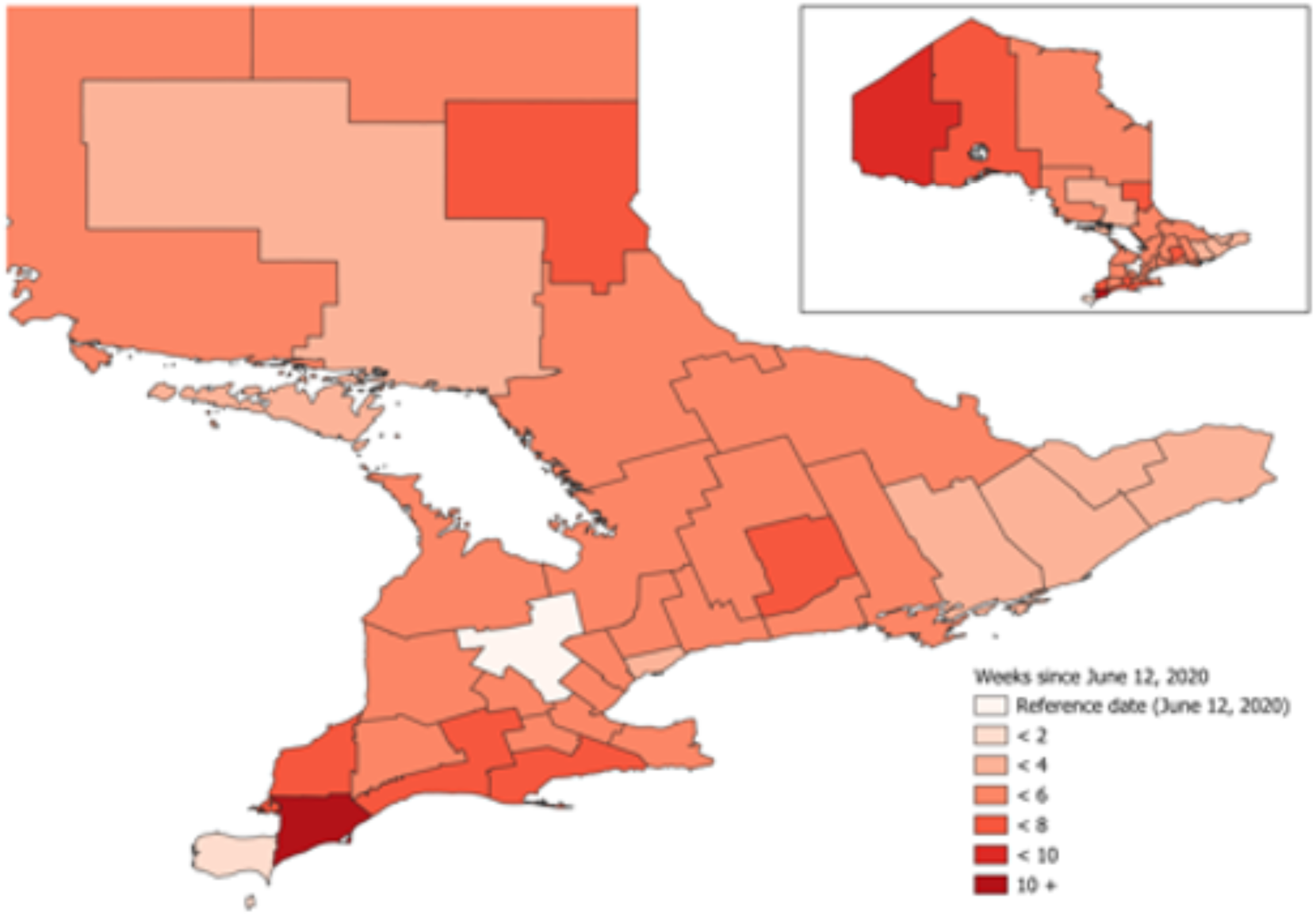
Timing of Indoor Mask Mandates in Ontario, Canada, 2020. Inset shows the entire province, while main body of the map is restricted to southern Ontario. Intensity of colour represents the lag since introduction of indoor mask mandates in the Wellington-Dufferin-Guelph public health unit on June 12, 2020. Northwestern health unit (inset, left side of map) and the Chatham-Kent health unit were the last public health units to introduce mask mandates, with Chatham-Kent’s mandate introduced on September 14, 2020.

We fit negative binomial regression models that included adjustment for time trends, age, female gender, public health unit and staged reopening for both reported cases and test-adjusted cases. For both models, the addition of masking significantly improved model fit (P<0.001 for likelihood ratio test for both models). For both dependent variables, indoor mask mandates were associated with a significant reduction in incidence (for reported cases, IRR 0.687; 95% CI 0.610 to 0.773; for test-adjusted cases IRR 0.512, 95% CI 0.467 to 0.561). The effect of masking was significantly stronger when test-adjusted cases were used as the dependent variable (P < 0.001 by Wald test).

We evaluated the robustness of these findings with alternate modeling approaches. For Model 2 and Model 3, masking was associated with reduced risk when reported cases were modeled, but these effects were not statistically significant (IRR 0.816, 95% CI 0.593 to 1.222 and IRR 0.576, 95% CI 0.240 to 1.385, respectively). By contrast, larger, and statistically significant, protective effects were seen with test-adjusted cases (IRR 0.494, 95% CI 0.358 to 0.681 and IRR 0.269, 95% CI 0.138 to 0.523, respectively). Results of all three models, and both outcome variables, are presented as mask mandate effectiveness estimates in **Table 1**.

**Table 1.** Estimated Effectiveness of Mask Mandates in Reducing SARS- CoV-2 Incidence, Ontario, Canada, 2020.

|  | **Reported Cases** | **Test-Adjusted Cases** |
| --- | --- | --- |
| **Model 1** | 31% (23-39%) | 49% (44-53%) |
| **Model 2** | 15% (-28% to 43%) | 51% (32-64%) |
| **Model 3** | 42% (-39% to 76%) | 73% (48-86%) |

**NOTE:** Model 1 is a negative binomial regression model with a log link and public health units treated as indicator variables; Model 2 is negative binomial panel data model with public health units treated as fixed effects; and Model 3 is generalized linear model a Poisson regression model with a log link and public health units treated as random effects.

We explored the possibility that the effects of masks might be modified by urbanicity (Greater Toronto/Hamilton Metropolitan Area vs. elsewhere), older age (60 or older), or gender by adding multiplicative interaction terms to our initial negative binomial model (Model 1). No significant interaction was seen between mask effect and older age (P = 0.366). However, the effect of mask mandates was significantly stronger within the province’s major urban area (Greater Toronto/Hamilton Area (GHTA)) than outside (P for interaction < 0.001; IRR within GTHA 0.370, 95% CI 0.332 to 0.413; IRR outside the GTHA 0.533, 95% CI 0.487 to 585). There was statistically significant interaction between gender and mask effect (P = 0.016), although differences in mask effect between females (IRR 0.496, 95% CI 0.450 to 0.546), and males (IRR 0.530, 95% CI 0.482 to 0.583), were small.

We estimated prevented fractions for masking during the period from June 12, 2020 to December 8, 2020 by comparing predictions from Model 1 to predictions from Model 1 with mask effect set to zero (**Figure 4**. With masking, Model 1 predicted 338,297 cases during this period (as compared to our test-adjusted estimate of 324,311 cases). When mask effect was set to zero, cases rose to 629,137, or 1.860 (95% CI 1.706 to 2.027) times higher than occurred with mask mandates. The prevented fraction due to mask mandates was estimated to be 46.2% (95% CI 41.4% to 50.7%). Based on age-specific hospitalization risk, intensive care admission risk, and case-fatality, as well as healthcare costs, we estimated that Ontario’s mask mandates prevented 3008 deaths, 9546 hospitalizations, 1879 hospital admissions, and the loss of 29,038 quality-adjusted life years. We estimate that the cost of premature deaths and healthcare usage averted through community mask mandates over this period had a value of approximately $CDN 610 million (**Appendix 1**).

**Figure 4.**
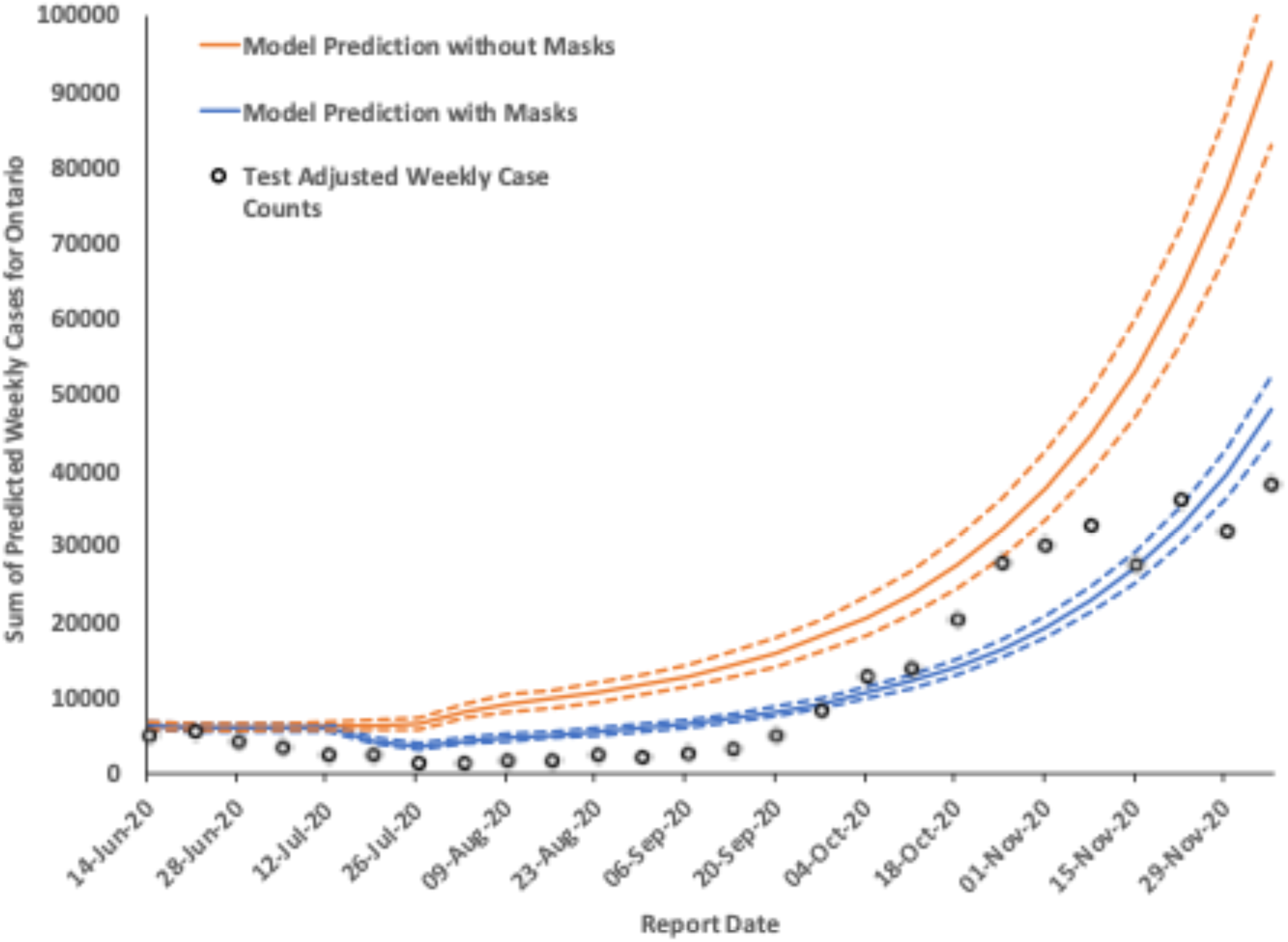
Model-Based Estimation of Mask Impact. Test-adjusted weekly case counts (circles) were used to fit a negative binomial regression model that included mask effects (predictions presented in the blue curve; dashed curves are 95% confidence intervals). Predictions from a negative binomial model with mask effects set to zero, but all other covariates identical, is shown as an orange curve (dashed lines are 95% confidence intervals). The gap between the two modeled curves is the estimated fraction of cases prevented by indoor mask mandates.

## Discussion

While the physical properties of masks and respirators reduce both production of infectious aerosols containing SARS-CoV-2 (25), and inhalation of infectious aerosols (26), the application of masks and respirators in indoor settings during the pandemic has been variable (27–30) and controversial (4, 5). Both real-world evidence and randomized trial evidence on the effects of community-level masking have been mixed in strength (6, 10, 31–36). While Ontario’s irregular, local introduction of indoor mask mandates established a quasi-experiment ideal for evaluation of mask effects, and earlier work suggests a modest reduction in SARS-CoV-2 incidence was associated with these mask mandates (10), we found that a large and robust effect of masks was seen once data were adjusted for under-testing in younger individuals and in males.

Our findings likely identify an important source of bias towards the null in the available literature on community masking effects. We find that the effects of masking are obscured by disproportionate testing of older individuals who are likely to present with more severe SARS-CoV-2 infection (37), and under-testing of younger individuals (children, teens, young men) who are less likely to undergo testing, and more likely to experience minimally symptomatic infection (38). Younger individuals are expected to contribute heavily to transmission dynamics, both because of density of social contacts (39), and (among younger men) attitudes towards risk (27). While this would be expected to be an important source of bias towards the null in observational studies (31, 32), it would also be a source of bias in randomized trials with differential identification of symptomatic infection by age such as (35), and indeed may partially explain the finding of greater mask effectiveness in older individuals in that study.

Other sources of bias towards the null in evaluations of community masking include imperfect compliance with mask mandates. Our finding of diminished effects of mask mandates in males may reflect apparent gender differences in compliance with public health measures more generally (27, 28, 30). However, the bidirectional effects of masking (8), which both protect wearers and those around them (by acting as “source control”) are also likely to bias towards the null, as reduced infectivity of mask users would have the effect of protecting non-users, thus narrowing the difference in risk between wearers and non-wearers. Our use of a quasi-experimental design with masking treated as a ubiquitous exposure would have reduced the impact of such a bias.

We also identified a significant difference in mask effectiveness in the province’s principal urban area (the Greater Toronto/Hamilton Area) as compared to less urban areas. Possible hypotheses about the mechanism underlying such an effect might include differences in density and contact patterns between urban and less urban areas, or differences in exposure to settings (e.g., subways or large indoor manufacturing facilities) where masks may have been particularly helpful in reducing transmission.

Our test adjustment serves as a means to correct for the fact that infection incidence during the SARS-CoV-2 pandemic was strongly determined by rates of clinical testing. The usual gold standard for identification of true infection incidence, as opposed to rates of case identification, is serology. However, in the context of SARS-CoV-2 in Canada, serological data have a number of important limitations, including performance of seroepidemiological studies in non-representative populations such as blood donors, the imperfect sensitivity and specificity of SARS-CoV-2 serological assays, and challenges in identifying repeated infections, as well as difficulty in differentiating infection from vaccination when anti-S antibody assays are used. Our methodology makes it possible to more accurately evaluate the impact of interventions on infection incidence even when serological data are limited in availability, accuracy or representativeness.

Like any epidemiological study ours has limitations. Key among these is the question of generalizability of mask effectiveness from Ontario to elsewhere in Canada, or to countries outside Canada. Furthermore, the effects we describe are observed at a time period when the ancestral strain of SARS-CoV-2 was circulating, and prior to the availability of SARS-CoV-2 vaccines and emergence of variants with increased infectivity. We are unlikely to have the opportunity to repeat this work in the current epidemiological context.

In summary, we find that adjustment for under-testing in younger age groups demonstrates that community mask mandates in Ontario, Canada were highly effective, and these effects were robust to different modeling approaches. Community masking mandates prevented substantial health and economic benefits for the province. Such mandates should be considered a potent tool for the management of future respiratory virus emergences.

## Data Availability

Aggregate data files needed for replication of results are available at https://figshare.com/articles/dataset/Data_for_Fisman_et_al_Test-Adjusted_Incidence_of_COVID-19_in_Ontario/14036528. The study was approved by the Research Ethics Board of the University of Toronto (protocol number 00044787).

---

**Appendix 1.**
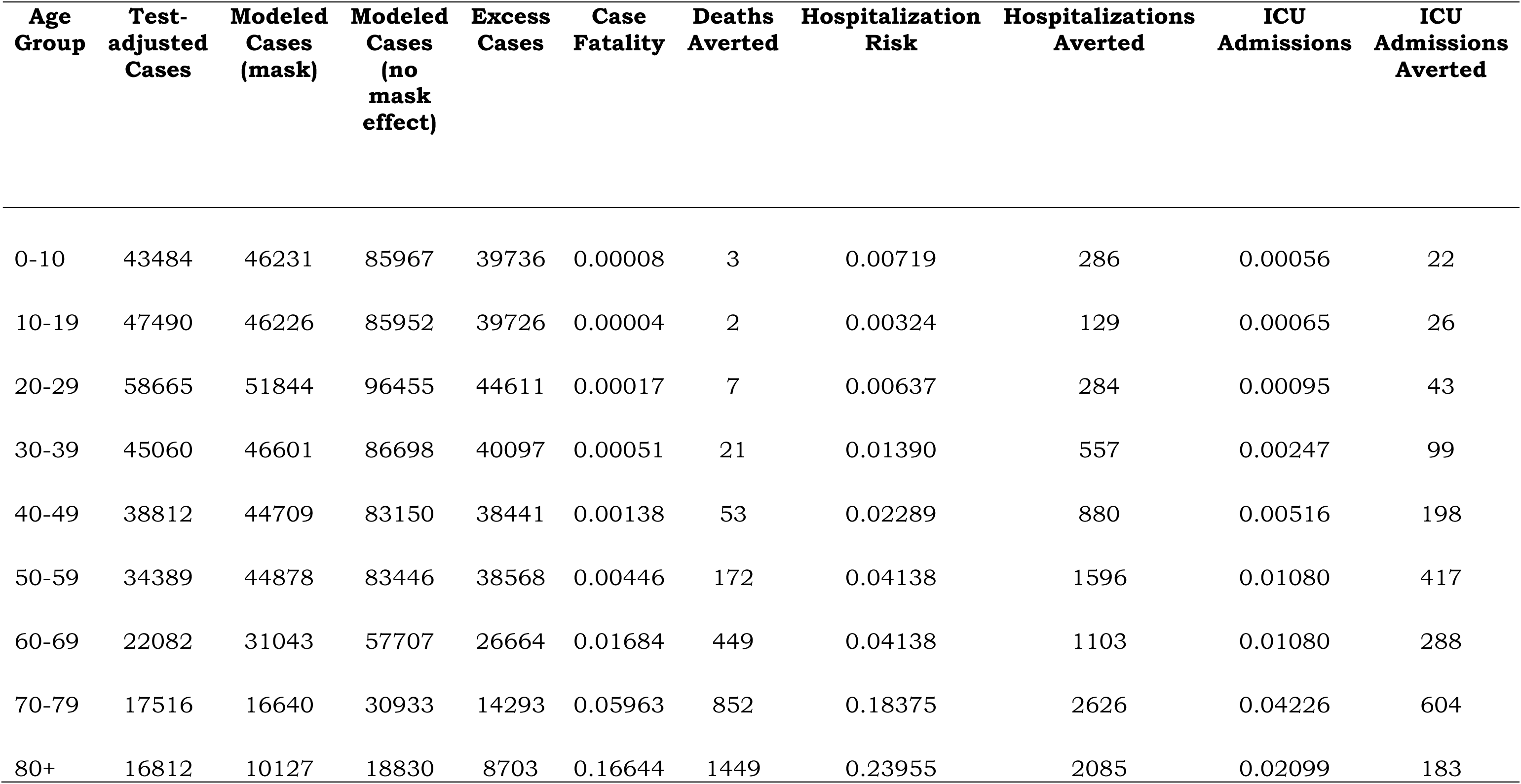
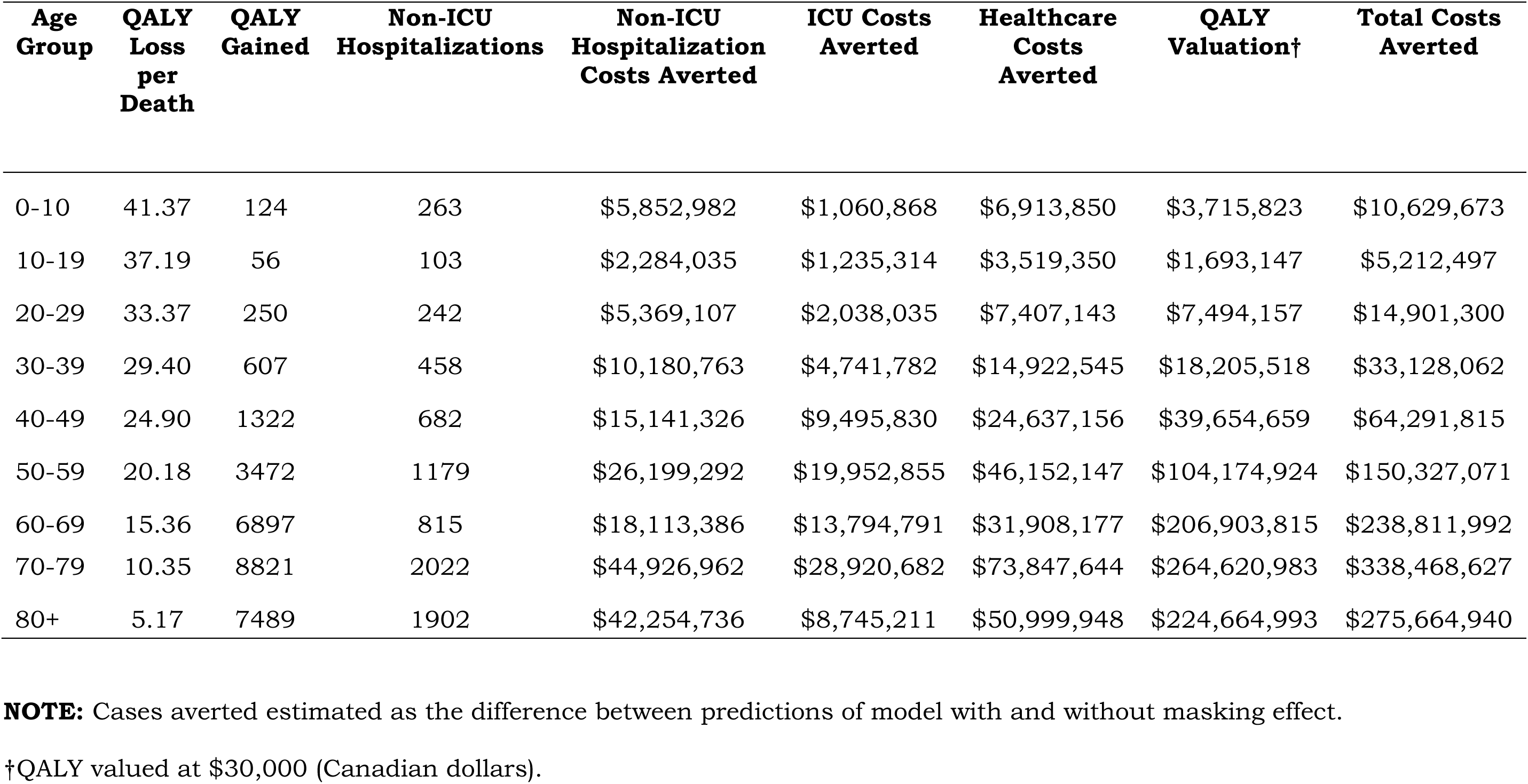
Estimated SARS-CoV-2-Related Health and Economic Consequences Averted through Community Masking Mandates, June 12-December 8, 2020.

| **Age Group** | **Test-adjusted Cases** | **Modeled Cases (mask)** | **Modeled Cases (no mask effect)** | **Excess Cases** | **Case Fatality** | **Deaths Averted** | **Hospitalization Risk** | **Hospitalizations Averted** | **ICU Admissions** | **ICU Admissions Averted** |
| --- | --- | --- | --- | --- | --- | --- | --- | --- | --- | --- |
| 0-10 | 43484 | 46231 | 85967 | 39736 | 0.00008 | 3 | 0.00719 | 286 | 0.00056 | 22 |
| 10-19 | 47490 | 46226 | 85952 | 39726 | 0.00004 | 2 | 0.00324 | 129 | 0.00065 | 26 |
| 20-29 | 58665 | 51844 | 96455 | 44611 | 0.00017 | 7 | 0.00637 | 284 | 0.00095 | 43 |
| 30-39 | 45060 | 46601 | 86698 | 40097 | 0.00051 | 21 | 0.01390 | 557 | 0.00247 | 99 |
| 40-49 | 38812 | 44709 | 83150 | 38441 | 0.00138 | 53 | 0.02289 | 880 | 0.00516 | 198 |
| 50-59 | 34389 | 44878 | 83446 | 38568 | 0.00446 | 172 | 0.04138 | 1596 | 0.01080 | 417 |
| 60-69 | 22082 | 31043 | 57707 | 26664 | 0.01684 | 449 | 0.04138 | 1103 | 0.01080 | 288 |
| 70-79 | 17516 | 16640 | 30933 | 14293 | 0.05963 | 852 | 0.18375 | 2626 | 0.04226 | 604 |
| 80+ | 16812 | 10127 | 18830 | 8703 | 0.16644 | 1449 | 0.23955 | 2085 | 0.02099 | 183 |

| Age Group | QALY Loss per Death | QALY Gained | Non-ICU Hospitalizations | Non-ICU Hospitalization Costs Averted | ICU Costs Averted | Healthcare Costs Averted | QALY Valuation† | Total Costs Averted |
| --- | --- | --- | --- | --- | --- | --- | --- | --- |
| 0-10 | 41.37 | 124 | 263 | \$5,852,982 | \$1,060,868 | \$6,913,850 | \$3,715,823 | \$10,629,673 |
| 10-19 | 37.19 | 56 | 103 | \$2,284,035 | \$1,235,314 | \$3,519,350 | \$1,693,147 | \$5,212,497 |
| 20-29 | 33.37 | 250 | 242 | \$5,369,107 | \$2,038,035 | \$7,407,143 | \$7,494,157 | \$14,901,300 |
| 30-39 | 29.40 | 607 | 458 | \$10,180,763 | \$4,741,782 | \$14,922,545 | \$18,205,518 | \$33,128,062 |
| 40-49 | 24.90 | 1322 | 682 | \$15,141,326 | \$9,495,830 | \$24,637,156 | \$39,654,659 | \$64,291,815 |
| 50-59 | 20.18 | 3472 | 1179 | \$26,199,292 | \$19,952,855 | \$46,152,147 | \$104,174,924 | \$150,327,071 |
| 60-69 | 15.36 | 6897 | 815 | \$18,113,386 | \$13,794,791 | \$31,908,177 | \$206,903,815 | \$238,811,992 |
| 70-79 | 10.35 | 8821 | 2022 | \$44,926,962 | \$28,920,682 | \$73,847,644 | \$264,620,983 | \$338,468,627 |
| 80+ | 5.17 | 7489 | 1902 | \$42,254,736 | \$8,745,211 | \$50,999,948 | \$224,664,993 | \$275,664,940 |

**NOTE:** Cases averted estimated as the difference between predictions of model with and without masking effect.
†QALY valued at \$30,000 (Canadian dollars).

